# Treadmill walking underestimates real-world walking spatiotemporal parameters and overestimates physiological demand across inclined terrain: implications for mobility assessment in older adults

**DOI:** 10.64898/2026.08.11.26360117

**Authors:** Keven Santamaria-Guzman, Tyrone Loria-Calderon, Mynor Rodriguez-Hernandez, Damaris C. Cifuentes, Silvia E. Campos-Vargas, Wendi H. Weimar, Ryan M. Babl, Yadrianna Acosta-Sojo, Kelly L. Thatcher, Jason R. Franz, David T. Redden, Brandon M. Peoples, Kenneth D. Harrison, Bria R. Smith, Francisco Siles-Canales, Jaimie A. Roper

## Abstract

**Purpose:** Treadmills (TM) are widely used for gait assessment in older adults (OA), yet their ecological validity across inclined terrain remains underexplored. This study compared spatiotemporal, physiological, and kinetic gait outcomes between TM and overground (OG) walking across flat, uphill, and downhill terrain in OA and younger adults (YA), and examined sensorimotor predictors of speed discrepancies.

**Methods:** Twenty-six OA (70 ± 6 years; 22 women) and 24 YA (26 ± 5 years; 7 women), none with prior TM experience, completed matched TM and OG trials across three terrain conditions. Self-selected TM speed was determined using a bidirectional protocol. The modified Clinical Test of Sensory Interaction in Balance quantified sensorimotor profiles. Mixed-design ANCOVAs and multiple regression examined condition, inclination, and group effects with sex as a covariate.

**Results:** TM speeds were consistently slower than OG across all conditions in both groups (Δ = −0.35 m/s, d = −1.67), with shorter stride length, lower cadence, and altered support phase timing; YA showed larger reductions and a greater shift toward double support than OA. Foot clearance at midswing was largely preserved across modalities. TM walking elicited higher heart rate and RPE despite slower speeds, most pronounced in OA uphill. Ground reaction forces and loading rates were substantially reduced on the TM. Sensorimotor profiles predicted the downhill speed discrepancy (R² = 0.49), with vestibular and somatosensory contributions as independent predictors alongside age group.

**Conclusion:** TM-derived speed, spatiotemporal, and physiological measures are not interchangeable with real-world ambulation data in OA across inclined terrain.

## Introduction

Treadmills (TM) are widely used in clinical and research settings for gait assessment, offering continuous data collection without spatial constraints and systematic control of walking speed and terrain inclination [1, 2]. Existing evidence supports broad biomechanical comparability between TM and overground (OG) walking on level surfaces in younger adults after appropriate familiarization [1, 3–10], though systematic differences in gait variability and spatiotemporal organization persist [3, 11].

Critical gaps limit the ecological validity of TM-based assessment for older adults (OA). First, TM-OG comparisons have almost exclusively examined level surfaces [6, 8, 9, 11–17], despite community ambulation routinely involving inclined terrain that imposes distinct demands on muscle activation, joint mechanics, metabolic responses, with direct implications for injury risk and bone health in older populations [18–21]. Second, while spatiotemporal gait parameters (e.g., stride length, cadence, gait speed) have been extensively used to characterize TM-OG differences on level surfaces [1, 3], whether terrain inclination modifies this spatiotemporal reorganization remains underexplored, a consequential gap given that inclined and declined walking imposes fundamentally different neuromuscular and mechanical demands [22–24].

Third, OA are substantially underrepresented in this literature relative to younger adults (YA; typically 20–40 years) [1, 17], despite age-related changes in sensorimotor integration, postural control, and neuromuscular function potentially producing fundamentally different locomotor adaptation strategies across both level and inclined terrain [25]. Fourth, whether individual sensorimotor profiles moderate differences in TM-OG walking remains entirely unexplored, despite its direct clinical relevance for fall risk screening and exercise prescription in OA.

The present study compared spatiotemporal, physiological, and kinetic gait outcomes between TM and OG walking across flat, inclined, and declined terrain in older and younger adults, and examined sensorimotor predictors of TM-OG discrepancies.

## Methods

### Participants

Twenty-six community-dwelling OA (22 women, 4 men; age 70 ± 6 years; mass 70.6 ± 10.5kg; height 161.1 ± 9.1 cm) and 24 YA (7 women, 17 men; age 26 ± 5 years; mass 72.4 ± 15.4 kg; height 166.8 ± 9.1 cm) were recruited from the University of Costa Rica. All walked independently without an assistive device and had no prior TM experience. Exclusion criteria included musculoskeletal, neurological, cardiovascular, respiratory, visual, vestibular, or cognitive impairments. The unequal sex distribution between groups was addressed by entering sex as a covariate in all analyses. All participants provided written informed consent.

### Instrumentation

TM walking was conducted on a Spirit CT800 TM (Spirit Fitness, Jonesboro, AR, USA). Spatiotemporal gait parameters were captured using six APDM Opal wireless inertial measurement units (IMUs; APDM Wearable Technologies, Portland, OR, USA) positioned bilaterally on the feet and wrists, with additional units on the lumbar spine (L5) and sternum, sampling at 128 Hz. Plantar pressure was recorded using pressure-sensing insoles (Loadsol, Novel Electronics, St. Paul, MN, USA) to estimate peak vertical ground reaction force (vGRF; normalized to body weight [BW]) and loading rate. Heart rate (HR) was monitored continuously via a wireless chest-strap monitor (Polar H9, Polar Electro, Kempele, Finland). All devices were calibrated per manufacturer specifications prior to data collection.

### Overground Protocol

Participants completed walking trials across three terrain conditions: a flat cement surface, and an outdoor ramp (mean slope 5°; 80 m total length) traversed uphill and downhill with a gradient conforming to accessibility specifications [26, 27]. This is the maximum slope permitted for built environments, ensuring ecological relevance to terrain routinely encountered by OA. Each trial lasted approximately two minutes, capturing a minimum of 30 complete strides [28]. Participants were instructed to walk at their “typical comfortable walking speed” without additional cueing [29]. A five-minute seated rest between trials allowed HR recovery to the resting baseline established at the start of the session. RPE was recorded immediately following each trial [30].

### Treadmill Protocol

On the same testing day, participants completed TM trials at matched inclination conditions. Self-selected walking speed was determined using a bidirectional protocol: speed began at 0.5 m/s and increased by 0.05 m/s every two seconds until participants verbally indicated their comfortable pace (incremental phase), then began at 2.5 m/s and decreased by 0.05 m/s every two seconds until comfortable pace was again identified (decremental phase). The average of both speeds defined each participant’s individualized walking velocity. Participants walked at this speed for approximately two minutes per condition, while HR and RPE were recorded. A five-minute seated rest separated condition. All six conditions were presented in a randomized order.

### Balance Assessment

The modified Clinical Test of Sensory Interaction in Balance (mCTSIB) was administered [31]. Sensory contribution scores were derived from the 95% ellipse sway area recorded by the lumbar IMU: Vision = eyes closed firm/eyes open firm; Somatosensory = eyes open foam/eyes open firm; Vestibular = eyes closed foam/eyes open firm. Higher scores indicate greater reliance on that sensory system for postural control [32].

### Data Processing

From all captured strides per trial, 30 consecutive strides were extracted, excluding initiation and cessation phases to ensure stable gait patterns. Spatial-temporal variables were calculated as the mean across these strides. Peak vGRF was defined as the absolute maximum force during the stance phase (normalized to BW), regardless of whether it occurred during the loading response or push-off; loading rate was calculated as the slope of the force–time curve from initial contact to peak force, averaged across 30 strides.

### Statistical Analysis

Mixed-design ANCOVAs examined the effects of walking condition (TM vs. OG), terrain inclination (flat, incline, decline), and age group (OA vs. YA) on all outcomes, with sex as a covariate. Sphericity was evaluated using Mauchly’s test, with Greenhouse-Geisser or Huynh-Feldt corrections applied where violated. Effect sizes are reported as partial eta squared (η²); significant effects were followed up with Bonferroni-corrected pairwise comparisons, with Cohen’s d reported for key contrasts.

Sensorimotor predictors of TM-OG discrepancies were examined using three separate multiple regression analyses (one per terrain inclination), with mCTSIB-derived sensory scores, age group, and sex as simultaneous predictors. Between-group sensory score differences were examined using Mann-Whitney U tests with rank biserial correlation (r), given significant normality violations (Shapiro-Wilk, all p < .001). All analyses were performed in JASP version 0.96.0 (JASP Team, 2026). Post-hoc power analysis indicated N = 50 achieved >99% power for the primary condition effect (η² = 0.685, f = 1.475, α = .05).

## Results

Descriptive statistics for all outcome variables are presented in Table 1. One participant was excluded from regression analyses based on standardized residual and Cook’s distance exceeding conventional thresholds, yielding a regression sample of N = 49. Sex did not reach significance as a covariate in any ANCOVA (all p > .31) and is not discussed further.

**Table 1.** Descriptive statistics for all outcome variables by age group, walking condition, and terrain inclination.

| Outcome | Inclination | OA Treadmill | OA Overground | YA Treadmill | YA Overground |
| --- | --- | --- | --- | --- | --- |
| | | Mean ± SD | Mean ± SD | Mean ± SD | Mean ± SD |
| Gait Speed<br>(m/s) | Flat | 1.040 ± 0.201 | 1.356 ± 0.154 | 0.869 ± 0.185 | 1.265 ± 0.268 |
| | Incline | 0.935 ± 0.196 | 1.233 ± 0.155 | 0.845 ± 0.195 | 1.287 ± 0.257 |
| | Decline | 1.059 ± 0.216 | 1.300 ± 0.175 | 0.883 ± 0.187 | 1.291 ± 0.257 |
| Stride Length<br>(m) | Flat | 1.109 ± 0.217 | 1.336 ± 0.133 | 1.054 ± 0.135 | 1.357 ± 0.172 |
| | Incline | 1.041 ± 0.211 | 1.276 ± 0.137 | 1.057 ± 0.144 | 1.401 ± 0.162 |
| | Decline | 1.121 ± 0.229 | 1.268 ± 0.157 | 1.056 ± 0.14 | 1.366 ± 0.179 |
| Cadence<br>(steps/min) | Flat | 113.1 ± 12.43 | 122.4 ± 7.85 | 98.35 ± 12.63 | 110.8 ± 11.95 |
| | Incline | 108.6 ± 13.37 | 115.7 ± 6.90 | 95.28 ± 12.89 | 109.5 ± 12.54 |
| | Decline | 114.1 ± 14.22 | 122.7 ± 5.63 | 99.82 ± 11.20 | 112.5 ± 10.71 |
| Double<br>Support<br>(%GCT) | Flat | 17.10 ± 4.73 | 15.87 ± 2.72 | 23.18 ± 3.29 | 18.77 ± 3.49 |
| | Incline | 18.34 ± 5.06 | 15.75 ± 3.03 | 23.33 ± 3.48 | 17.51 ± 3.80 |
| | Decline | 17.41 ± 4.35 | 17.91 ± 3.25 | 22.86 ± 3.15 | 18.72 ± 3.44 |
| Single Support<br>(%GCT) | Flat | 41.46 ± 2.39 | 42.02 ± 1.31 | 38.42 ± 1.65 | 40.61 ± 1.74 |
| | Incline | 40.76 ± 2.39 | 42.12 ± 1.53 | 38.33 ± 1.74 | 41.18 ± 1.79 |
| | Decline | 41.48 ± 2.33 | 41.04 ± 1.57 | 38.57 ± 1.58 | 40.61 ± 1.65 |
| Elevation at<br>Midswing<br>(cm) | Flat | 2.714 ± 1.194 | 2.156 ± 0.7603 | 1.811 ± 0.8524 | 1.269 ± 0.7091 |
| | Incline | 3.608 ± 1.482 | 3.377 ± 0.7169 | 2.028 ± 0.9742 | 1.961 ± 0.7936 |
| | Decline | 1.93 ± 1.256 | 1.933 ± 0.8066 | 1.727 ± 0.7955 | 1.542 ± 0.5892 |
| Heart Rate<br>(bpm) | Flat | 111.4 ± 14.65 | 100.1 ± 12.58 | 97.08 ± 9.84 | 99.58 ± 10.34 |
| | Incline | 118.3 ± 13.84 | 104.8 ± 12.08 | 103.8 ± 9.07 | 93.42 ± 10.37 |
| | Decline | 105.7 ± 14.54 | 99.08 ± 12.55 | 94.67 ± 10.78 | 92.50 ± 10.58 |
| RPE 6-20<br>(Score) | Flat | 9.12 ± 2.47 | 8.27 ± 1.93 | 7.17 ± 1.34 | 7.04 ± 1.43 |
| | Incline | 10.08 ± 2.58 | 8.08 ± 1.92 | 8.38 ± 2.00 | 7.04 ± 1.40 |
| | Decline | 8.81 ± 2.28 | 8.00 ± 2.06 | 7.38 ± 1.47 | 6.75 ± 0.99 |
| Peak vGRF<br>(N) | Flat | 838.3 ± 105.2 | 986.6 ± 106.1 | 848.7 ± 103.7 | 1123 ± 218.8 |
| | Incline | 854.1 ± 97.96 | 952.9 ± 106.9 | 859.1 ± 106.2 | 1047 ± 177.9 |
| | Decline | 826.2 ± 116.0 | 1072 ± 162.4 | 855.1 ± 114.4 | 1143 ± 240.6 |
| Loading Rate<br>(N/ms) | Flat | 5.69 ± 2.39 | 9.56 ± 2.68 | 6.04 ± 2.01 | 10.03 ± 3.65 |
| | Incline | 5.27 ± 2.28 | 7.35 ± 2.59 | 5.48 ± 2.10 | 9.42 ± 2.85 |
| | Decline | 6.22 ± 2.15 | 10.45 ± 2.51 | 5.41 ± 1.65 | 11.62 ± 4.19 |

Notes: OA = Older Adult; YA = Younger Adults.

### Spatiotemporal

There were differences between TM and OG in gait speed for both OA (t = −7.181, p < .001, d = −1.343) and YA (t = −11.421, p < .001, d = −1.992) (Figure 1. A), and across all inclinations: Flat (t = −12.757, p < .001, d = −1.711), Incline (t = −12.981, p < .001, d = −1.741), and Decline (t = −11.567, p < .001, d = −1.551). Both age groups walked slower on the TM across all inclinations compared to OG (Figure 1. B).

**Figure 1.**
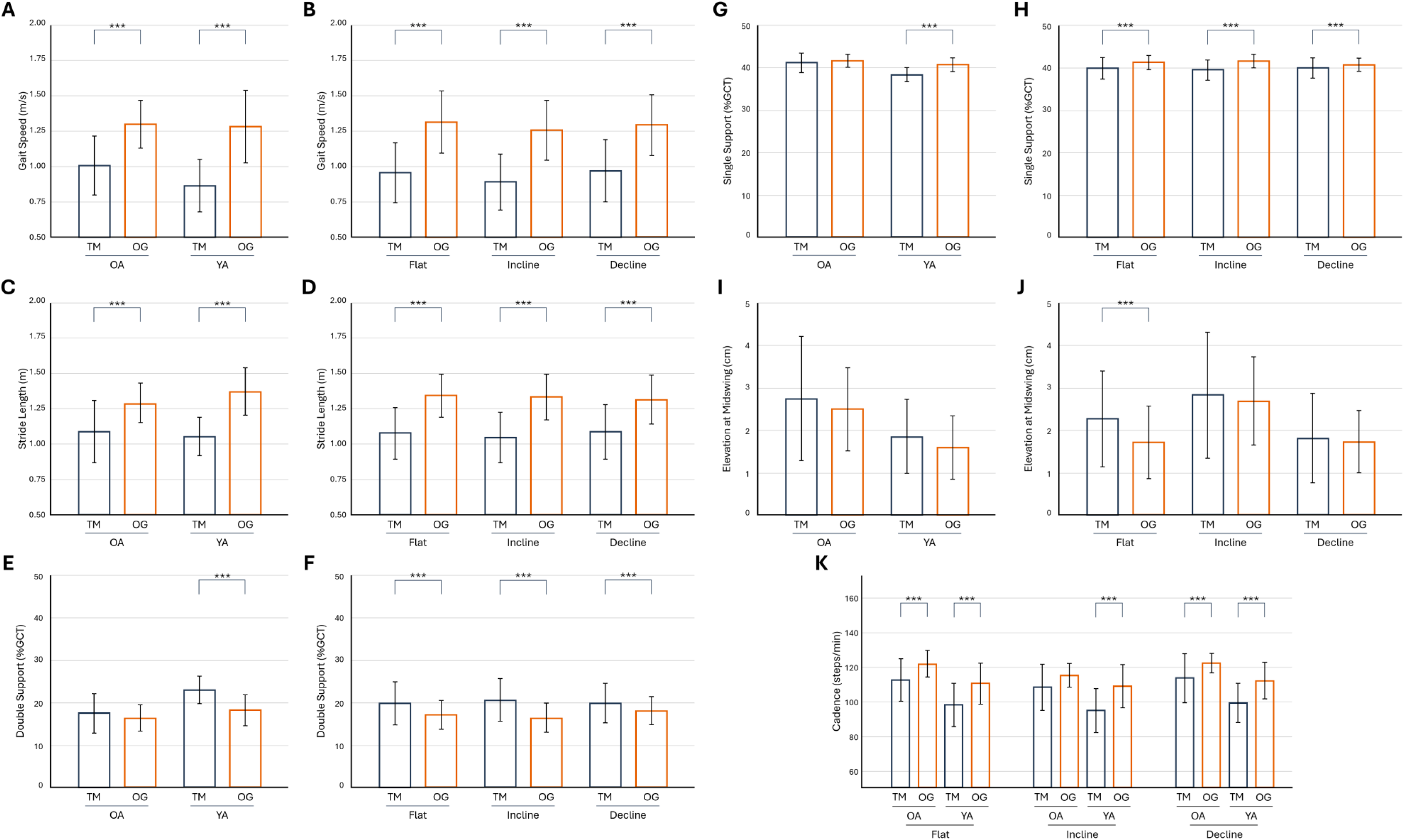
Spatiotemporal gait parameters during treadmill (TM) and overground (OG) walking by age group and terrain inclination. Notes: (A, B) Gait speed (m/s) by group (OA, YA) and by inclination (Flat, Incline, Decline). (C, D) Stride length (m) by group and inclination. (E, F) Double support (%GCT) by group and inclination. (G, H) Single support (%GCT) by group and inclination. (I, J) Elevation at midswing (cm) by group and inclination. (K) Cadence (steps/min) by inclination and group combined. Bars represent group means; error bars represent ± SD. Asterisks denote statistically significant pairwise comparisons (*: p < 0.05, **: p < 0.01, ***: p < 0.001).

The same pattern emerged for stride length, with differences between TM and OG in OA (t = −5.949, p < .001, d = −1.155) and YA (t = −10.332, p < .001, d = −1.870) (Figure 1. C), and across Flat (t = −10.764, p < .001, d = −1.525), Incline (t = −11.926, p < .001, d = −1.689), and Decline (t = −10.176, p < .001, d = −1.323); both groups showed shorter strides on the TM across all inclinations (Figure 1. D).

For cadence, differences emerged only within the triple interaction. OA showed lower TM cadence at Flat (t = −4.193, p = .008, d = −0.952) and Decline (t = −4.310, p = .006, d = −0.916), while YA showed lower TM cadence at Flat (t = −5.479, p < .001, d = −1.765), Incline (t = −6.136, p < .001, d = −1.355), and Decline (t = −5.825, p < .001, d = −1.155), indicating a significant cadence reduction on a TM and YA showed a consistently in all inclinations (Figure 1. K).

Double support time increase on a TM compared with OG in YA only (t = 7.174, p < .001, d = 1.320) (Figure 1. E), with no significant TM-OG difference in OA. And across all inclinations: Flat (t = 5.106, p < .001, d = 0.789), Incline (t = 7.280, p < .001, d = 1.162), and Decline (t = 3.681, p = .009, d = 0.463) (Figure 1. F). Single support showed decrement on a TM compared with OG in YA only (t = −7.322, p < .001, d = −1.266) (Figure 1. G) and across Flat (t = −5.063, p < .001, d = −0.746), Incline (t = −7.769, p < .001, d = −1.115), and Decline (t = −3.809, p = .006, d = −0.441), confirming that YA, but not OA, reorganized support phase timing on the TM across all terrain (Figure 1. H).

As a secondary outcome, midswing elevation did not differ between groups in any condition (Figure 1I). Condition differences were observed only during flat walking (t = 4.495, p < .001, d = 0.622), with no differences during incline or decline (Figure 1. J). This suggests that foot clearance was maintained across TM and OG walking on inclined and declined terrain.

### Physiological Demand

Within the three-way interaction, OA showed higher HR on the TM than OG at Flat (t = 5.687, p < .001, d = 0.974), Incline (t = 6.129, p < .001, d = 1.049), and Decline (t = 3.680, p = .024, d = 0.630); OA also exceeded YA on the TM at Flat (t = 4.015, p = .010, d = 1.373) and YA exceeded OA on OG at Incline (t = 5.596, p < .001, d = 0.902), while YA showed a TM-OG difference only at Incline (t = 5.849, p < .001, d = 1.959) (Figure 2. A). This indicates HR was consistently elevated on the TM relative to OG in OA across all terrain, while in YA the elevation was restricted to incline walking. For RPE, there were differences between TM and OG at Incline (t = 7.426, p < .001, d = 0.865) and Decline (t = 3.069, p = .039, d = 0.357) (Figure 2. B), indicating higher perceived exertion on the TM during inclined and declined walking.

**Figure 2.**
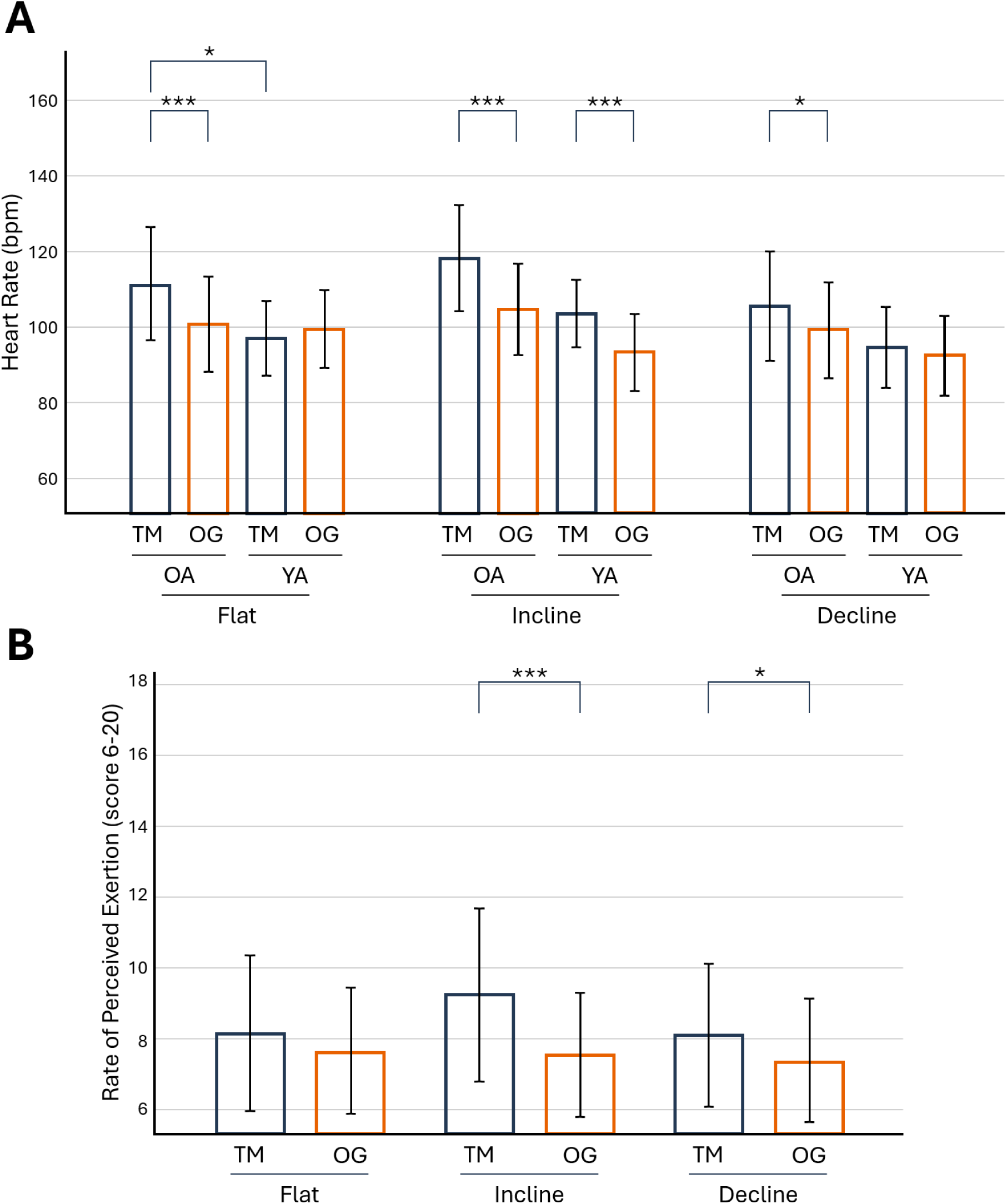
Heart rate and rating of perceived exertion during treadmill and overground walking across terrain inclinations. Notes: (A) Heart rate (bpm) by condition, inclination, and age group (OA, YA); bars shown in order OA-TM, OA-OG, YA-TM, YA-OG within each terrain cluster. (B) Rating of Perceived Exertion (RPE, 6-20 scale) by condition and inclination. Error bars represent ± SD. Asterisks denote statistically significant pairwise comparisons (*: p < 0.05, **: p < 0.01, ***: p < 0.001).

### Ground Reaction Kinetics

Peak vGRF differed between TM and OG in both OA (t = −6.598, p < .001, d = −1.138) and YA (t = −11.181, p < .001, d = −1.692), and across all inclinations: Flat (t = −9.444, p < .001, d = - 1.416), Incline (t = −6.778, p < .001, d = −1.017), and Decline (t = −12.078, p < .001, d = −1.812) (Figure 3. A). This indicates peak vGRF was consistently higher on OG than TM across both groups and all terrain. Loading rate showed the same pattern, differing between TM and OG in both OA (t = −5.164, p < .001, d = −1.149) and YA (t = −9.231, p < .001, d = −1.823) (Figure 3. B), indicating higher loading rates on OG than TM in both groups.

**Figure 3.**
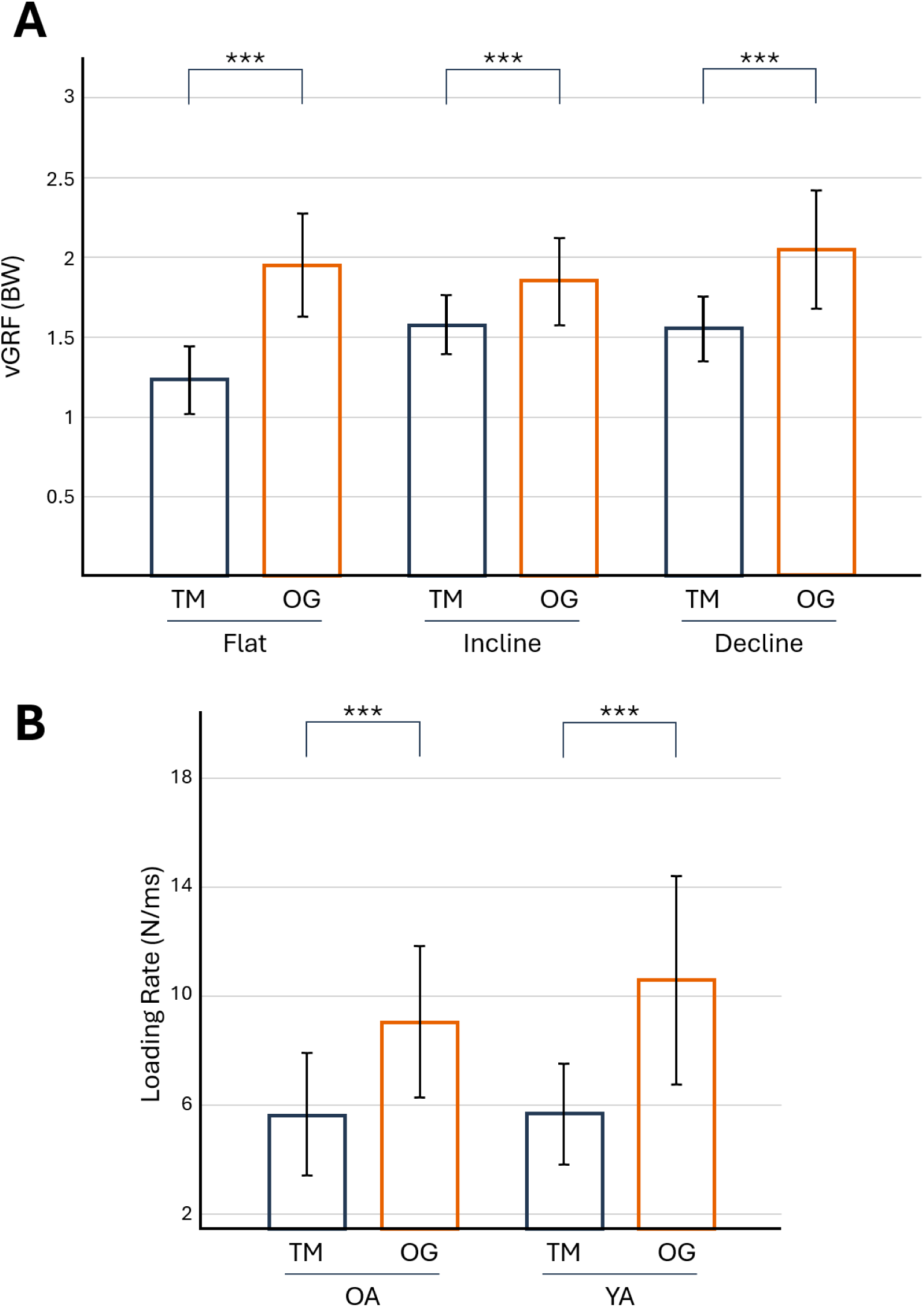
Peak vertical ground reaction force and loading rate during treadmill (TM) and overground (OG) walking. Notes: (A) Peak vertical ground reaction force (vGRF) normalized to BW by condition and terrain inclination, collapsed across age group. (B) Loading rate (N/ms) by condition and age group (OA, YA), collapsed across terrain. Error bars represent ± SD. Asterisks denote statistically significant pairwise comparisons (*: p < 0.05, **: p < 0.01, ***: p < 0.001).

### Sensorimotor Profiles and Speed Discrepancy

OA and YA did not differ significantly on any mCTSIB sensory domain (Vision: U = 356.5, p = .262, r = −0.188; Somatosensory: U = 245.0, p = .275, r = 0.183; Vestibular: U = 260.0, p = .429, r = 0.133; Mann-Whitney tests). The flat speed discrepancy model was not significant (F(5,42) = 1.522, p = .204, R² = 0.153). The incline model reached significance (F(5,42) = 2.905, p = .024, R² = 0.257), with group as the sole significant predictor (B = 0.152, p = .040). The decline model explained the most variance (F(5,42) = 8.117, p < .001, R² = 0.491, adjusted R² = 0.431), with vestibular CTSIB as the strongest positive predictor (B = 0.018, p < .001), somatosensory CTSIB as a significant negative predictor (B = −0.027, p = .020), and group as an independent predictor (B = 0.140, p = .012); vision and sex were not significant (both p > .20).

## Discussion

The most consistent finding was a large, systematic TM-OG walking speed discrepancy of approximately 0.35 m/s that persisted across flat, inclined, and declined terrain in both groups, accompanied by a coordinated reorganization of spatiotemporal gait pattern. Stride length and cadence were both reduced on the TM relative to OG across all inclinations, with YA showing larger reductions than OA, indicating the slower TM speed reflected a contraction of the entire gait cycle rather than a single compensatory adjustment; this is contrary to most studies analyzing spatiotemporal parameters between TM and OG [2, 3].

This reorganization extended to support phase timing; YA, but not OA, increased double support time and correspondingly decreased single support time on the TM across all terrain, a pattern consistent with a stability-prioritizing strategy adopted preferentially by younger adults when confronted with the TM’s external belt constraint [33, 34]. The absence of a comparable support-phase shift in OA may reflect a ceiling effect, as OA already operate with proportionally greater double support time during OG walking [13, 25] and may have limited additional margin to further prioritize stability on the TM.

Foot clearance at midswing, examined as a secondary safety-relevant outcome, was largely preserved across modalities, differing between TM and OG only on flat terrain; OA consistently elevated the foot higher than YA during incline walking regardless of modality, suggesting that age-related clearance strategy is terrain-driven rather than modality-driven. Together, these findings indicate that TM walking does not simply slow gait uniformly but reorganizes its underlying spatiotemporal structure in an age-dependent manner, with younger adults adopting a more conservative, stability-oriented pattern than older adults. Despite selecting slower TM walking speeds and a more conservative spatiotemporal gait pattern, participants exhibited significantly higher heart rate and RPE during TM than OG walking across all inclinations, a paradox that directly challenges the assumption of physiological equivalence between modalities [35, 36]. The three-way interaction for heart rate revealed that OA TM incline walking produced the highest cardiovascular response across all conditions, substantially exceeding both OA OG incline and YA TM incline responses, while YA showed an elevated TM-OG difference restricted to incline terrain.

The motorized belt may impose a continuous external locomotor demand that constrains the natural stride-to-stride adjustments characteristic of OG walking, increasing neuromuscular co-activation and metabolic cost independently of speed, compounded by elevated sympathetic arousal from the perceptual novelty of TM locomotion [2, 37, 38]. These findings carry two opposing clinical implications: heart rate or RPE targets calibrated on the TM may cause clinicians to underestimate community ambulation capacity [36], while TM training may not adequately replicate the neuromuscular and cardiovascular demands of real-world walking, potentially limiting transfer to community ambulation.

TM walking produced substantially lower peak vGRF and loading rates than OG walking across all inclinations in both groups. These differences require cautious interpretation, as walking speed is a primary determinant of ground reaction force magnitude; the observed kinetic differences likely reflect surface mechanical properties combined with speed-dependent biomechanical adaptation rather than modality alone [14, 39]. Belt compliance, where it contributes to force attenuation, is specific to consumer-grade and rehabilitation TMs with compliant decks, an effect absent in rigid-deck instrumented systems. The reduced musculoskeletal loading during TM walking may benefit populations with painful loaded joints [6, 14, 39]; however, the correspondingly reduced mechanical stimulus may limit bone density adaptations that depend on adequate impact loading.

The regression analyses revealed a terrain-specific pattern of sensorimotor prediction. The decline model explained the largest proportion of variance in the TM-OG speed discrepancy (R² = 0.491), with vestibular reliance as the strongest positive predictor and somatosensory reliance as a significant negative predictor, independent of age group and sex. Greater proprioceptive reliance may aid recalibration of locomotor speed to the TM belt via ongoing somatosensory input [40], while greater vestibular reliance may confer vulnerability, consistent with evidence that motorized belts disrupt multisensory integration strategies that normally regulate postural orientation during real-world decline walking [32].

The incline model also reached significance, driven primarily by age group membership rather than individual sensory scores, consistent with the hypothesis that YA adopt more conservative TM speed selection in a novel locomotor environment while maintaining greater OG incline speeds [21, 24]. These patterns carry direct clinical relevance: individuals with compromised somatosensory function, including those with peripheral neuropathy, radiculopathy, or post-surgical sequelae, may be particularly susceptible to large TM-OG speed discrepancies on inclined terrain, precisely the population most commonly referred for TM-based rehabilitation. Brief mCTSIB screening prior to inclined TM use may therefore help clinicians identify those most at risk of assessment error.

Several limitations warrant consideration. The unequal sex distribution between groups was addressed statistically by entering sex as a covariate, but residual confounding cannot be excluded. A single ramp gradient of 5° does not capture the full range of community terrain gradients encountered during daily ambulation. The cross-sectional, single-session design precludes examination of familiarization effects or test-retest reliability, and the modest sample size limits power for smaller interaction effects.

### Conclusion

TM walking produced slower speeds, shorter strides, and lower cadence than OG walking across all terrains, paradoxically accompanied by greater cardiovascular and perceptual demands. This reorganization was age-dependent: YA shifted toward a more stability-oriented pattern, whereas OA showed minimal adjustment in the support phase. Foot clearance was largely preserved across modalities, indicating terrain, not surface, governs this fall-relevant behavior. Discrepancies were most pronounced in OA during uphill walking and varied with sensorimotor profile during downhill walking, in which vestibular and somatosensory contributions independently predicted the magnitude of mismatch. TM-derived measures should not be assumed to be interchangeable with real-world ambulation in older adults; terrain-specific familiarization, sensorimotor screening, and modality-specific norms should guide TM-based assessment in this population.

## Data availability

The dataset used and analyzed during the current study is available from the corresponding author on reasonable request.

## Acknowledgements

The authors would like to thank the participants of the Comprehensive Program for Older Adults at the University of Costa Rica for their time and willingness to participate in this study.

## Declaration of generative AI and AI-assisted technologies in the manuscript preparation process

During the preparation of this work the authors used Claude (Anthropic) in order to refine the clarity, grammar, and flow of the written manuscript. All scientific content, including research design, methodology, and interpretation, was developed by the authors. The AI tool served only as an editing assistant to improve readability. After using this tool, the authors reviewed and edited the content as needed and take full responsibility for the content of the published article.

